# Combining models to generate consensus medium-term projections of hospital admissions, occupancy and deaths relating to COVID-19 in England

**DOI:** 10.1101/2023.11.06.23298026

**Authors:** Harrison Manley, Thomas Bayley, Gabriel Danelian, Lucy Burton, Thomas Finnie, André Charlett, Nicholas A Watkins, Paul Birrel, Daniela De Angelis, Matt Keeling, Sebastian Funk, Graham Medley, Lorenzo Pellis, Marc Baguelin, Graeme J Ackland, Johanna Hutchinson, Steven Riley, Jasmina Panovska-Griffiths

## Abstract

Mathematical modelling has played an important role in offering informed advice during the COVID-19 pandemic. In England, a cross government and academia collaboration generated Medium-Term Projections (MTPs) of possible epidemic trajectories over the future 4-6 weeks from a collection of epidemiological models.In this paper we outline this collaborative modelling approach and evaluate the accuracy of the combined and individual model projections against the data over the period November 2021-December 2022 when various Omicron subvariants were spreading across England. Using a number of statistical methods, we quantify the predictive performance of the model projections for both the combined and individual MTPs, by evaluating the point and probabilistic accuracy. Our results illustrate that the combined MTPs, produced from an ensemble of heterogeneous epidemiological models, were a closer fit to the data than the individual models during the periods of epidemic growth or decline, with the 90% confidence intervals widest around the epidemic peaks. We also show that the combined MTPs increase the robustness and reduce the biases associated with a single model projection. Learning from our experience of ensemble modelling during the COVID-19 epidemic, our findings highlight the importance of developing cross-institutional multi-model infectious disease hubs for future outbreak control.

## 1 Introduction

Mathematical modelling has played an important role in offering scientific advice to policy makers at important junctions over the COVID-19 pandemic. Modelling allows information from data and epidemiological theory to be used in a transparent and rigorous technical framework, within which current epidemic trajectories can be assessed and projections of future behaviours can be made. Epidemiological estimates of how a virus may spread in the future along with uncertainties and limitations surrounding these estimates are useful tools for future policy planning.

Over the COVID-19 epidemic a number of models have been developed and used at pace. Their main power is in populating the technical framework with data and generating (probabilistic) projections of possible futures. While fitting to historic data and processes gives tight constraints on the models’ behaviour in the past, projecting trends into the future can be uncertain and dependant on the assumptions of the models about future events such as schools opening/closing, level of social mixing or the intrinsic properties of the circulating virus which enable it to escape imposed intervention and vaccination mitigation strategies. Specifically, when modelling the future, modellers are faced with complex and uncertain scenarios. The decision-making process relies on them making assumptions grounded on what may happen in the future and quantifying the likelihood of those potential outcomes[1, 2].

Ensemble modelling is a quantitative method that combines information from multiple individual models to generate a combined or consensus outcome. It is a common practise in Climate Science [3, 4, 5], Economics [6] and weather forecasting [7, 8]. Similarly to climate modelling, uncertainties in epidemic model outputs arise from uncertainties in initial conditions, recorded observations, model assumptions and model structure. Since different models have different underlying assumptions and hence project different possible futures, aggregating and combining results from a number of models may mitigate some of the uncertainty of the possible futures. There are also uncertainties in the model parameters and structural uncertainties resulting from the fact that some processes in the modelled system are not fully understood, or are impossible to model completely due to computational constraints. Using a model ensemble can help to characterise the overall uncertainty in a system [9]. Furthermore, averaging the outputs of multiple model ensembles has been shown numerous times to compare more favourably with observations and yield better projections than a single model [10].

Combining epidemiological models to generate aggregated model outcomes pre-COVID-19 was applied to modelling HIV [11], influenza [12] and Ebola [13, 14] transmission. However, this has developed rapidly over 2020-2022 with a number of countries setting up modelling hubs to ensemble model the COVID-19 epidemic [15, 16, 17].

In the UK such a modelling hub was formed as a formal collaboration between the Department of Health and Social Care (DHSC) advisory committee on pandemic modelling (Scientific Pandemic Influenza Group on Modelling - Operational (SPI-M-O)) and the UKHSA Epidemiological Ensemble group (UKHSA Epi-Ensemble). Since early 2021 this modelling hub has provided the UK government with weekly Medium-Term Projections (MTPs) as a combined epidemic trajectory estimate from a number of epidemiological models (having taken over from SPI-M-O, who produced the MTPs before this in 2020). These comprise epidemic trajectories of hospital admissions, hospital bed occupancy and deaths over the future 4-6 weeks of the epidemic, and are generated from a set of epidemiological models maintained and run by members of SPI-M-O or the UKHSA Epi-Ensemble.

Production of the Medium Term projections began in late August 2020 and were initially generated by SPI-M-O on a weekly basis[18], to replace the short term forecasts that had been generated up to that point several times a week [19]. With the development of the SPI-M-O and UKHSA Epi-Ensemble modelling hub, the responsibility to produce MTPs was transferred to the UKHSA Epi-Ensemble in early 2021. As part of the collaboration, modelling teams within SPI-M-O and UKHSA Epi-Ensemble were asked to produce projections under an explicit assumption of no changes other than population immunity[20]. This was done to allow policy makers to assess the likely outcomes of the epidemic based on current policy interventions. Whilst the modelling teams were provided with expected vaccination rates and asked to incorporate known factors such as school term times, in general the MTPs were not planned to be forecasts. Therefore, evaluating them as such deviates from their original purpose. However, the analyses undertaken here give valuable retrospective insight into the capability of models used during a pandemic, which is useful for future pandemic preparedness planning. Furthermore, the time frame considered here almost entirely coincides with there being no legal COVID restrictions, so the differences between predictions and projections for the purposes of this analysis are minimal.

As we discussed in our previous paper [21], the value in getting a combined forecast from across models and datasets is not just in the weighted averaging of those estimates, but also in the formation of a community that are constantly discussing the outcomes, the modelling assumptions and the input data, identifying the drivers behind the differences across models’ outcomes when formulating the aggregated possible future projections.

Whilst the United Kingdom was not the only country to produce COVID-19 forecasts from a model ensemble, thorough descriptions and evaluations of the model ensembles used are currently lacking. For example, modellers in the United States, in conjunction with the Center for Disease Control (CDC), published ensemble forecasts using a wide variety of mathematical models [2, 22] forecasting new cases, hospitalisations and deaths at a national and state-level. However, the current evaluation of the US ensemble analysed only the performance of deaths forecasting over a small section of the pandemic between May 2020 and July 2020 [2]. As such, ensemble forecast performance on other policy relevant metrics such as hospital admissions and hospital occupancy has not been considered, and there is a lack of insight into how forecast performance may vary at different points of the COVID-19 epidemic.

In light of the above, this paper outlines how MTPs were generated for England using a previously established combination method [23] throughout the COVID-19 pandemic, as a collaboration across government and academia. We detail the approach of generating a single, combined consensus model projection from an ensemble of multiple epidemiological models applied to the English epidemic, and how they were combined to produce aggregated MTPs over time. We follow this with a detailed evaluation of the performance of the generated combined estimates over the period November 2021-December 2022, noting when the MTPs were better or worse at estimating the current epidemic status and projecting forward trajectories and exploring why.

## 2 Methodology

### 2.1 Models used to produce MTPs

A number of mathematical models have been developed, adapted and used throughout 2020-2022 to model the COVID-19 epidemic and produce MTPs in England. The models described in this paper broadly fall into three groups: population based (PBMs), agent based (ABMs) and data driven models (DDMs). Table A1 contains the models in the ensemble, with a description of their main characteristics.

### 2.2 Data Sources

Data sources are important in modelling - both in parameterising the models and in calibrating/validating them. In this ensemble, the models were informed by a range of different data sources, and were fit to specific data for the projections of each metric. In the data that was used, and therefore for the purposes of this paper, the metrics are defined as in table 1. This data was provided to the modellers via a secure transfer from internal sources. A summary of the data to which each model is fitted to is described in table A1.

**Table 1.** : The definitions of the metrics used from the data source.

| Metric | Data definition |
| --- | --- |
| Hospital admissions | Daily number of inpatients with a positive diagnosis of COVID-19 |
| Hospital bed occupancy | Daily number of hospital beds occupied with confirmed COVID-19 patients |
| Deaths | Daily number of people with laboratory-confirmed positive COVID-19 tests, who died within (equal to or less than) 28 days of the first positive specimen date. |

### 2.3 Combining model projections to generate a consensus MTP

Each of the calibrated epidemiological models described in table A1 were able to produce MTPs for one or more of: hospital admissions, hospital occupancy and deaths over the future 4-6 weeks.^1^ The results from the modelling groups were submitted as quantiles of the posterior predictive distribution of the model’s outcomes, with quantiles ranging from the 5th to 95th in increments of 5. Posterior predictive distributions were then estimated for each model as skewed-Normal distributions fitted to the submitted set of quantiles[23, 24].

To illustrate how these distributions were then combined, we can consider a list of epidemiological models = (*M*_1_, *M*_2_, …, *M*_*N*_ ), which are fitted to observed data y and generate projected data f, where the n_th_ model has a posterior predictive distribution *p*_*n*_(*f* | *y*), the model ensemble will then have a posterior predictive density of the form:

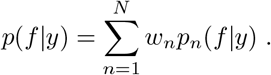

In the above, *w*_*k*_ ≥ 0 weighs the *k*_*th*_ model’s contribution to the ensemble, and 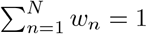.For the combined projections we used an equal weights combination method, setting 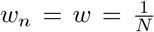 The visualisation of the projections was implemented using CrystalCast[25], a piece of software developed by the Defence Science and Technology Laboratory (DSTL).

### 2.4 Assessing model performance

The performance of each model can be assessed quantitatively by evaluating the point and probabilistic accuracy of the individual model projections. We examined the predictive performance for England for a range of forecast horizons from *t* ∈ [*t*_0_, *t*_0_ + 7] to *t* ∈ [*t*_0_, *t*_0_ + 28] to highlight how the accuracy changes for different projection lengths, where t_0_ is the date of the first day of the forecast. The methods we used to evaluate and compare the predictive accuracy of the models are detailed below.

#### 2.4.1 Mean Absolute Error (MAE)

The Absolute Error (AE) was evaluated for each point of the projections. For a given model this is the absolute difference between the projected median and the recorded data timeseries. The MAE then gives the mean of this value for a given projection, taken over the entire forecast horizon. The MAE is useful as it provides an intuitive, quantitative estimate of the forecast performance in the natural units of the data. To calculate this, we used the standard formula:

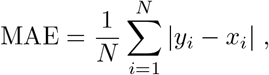

where *N* is the length of the forecast horizon in days, *y*_*i*_ is the central estimate of the projection and *x*_*i*_ is the recorded data on day *i*. We calculated the MAE for each published MTP for each metric, to examine the trend across the time period and observe how the forecast accuracy changes with successive Omicron variants.

#### 2.4.2 Weighted interval score

We also assessed the probabilistic accuracy using a scoring rule. There were many proper scoring rules at our disposal from the literature [26]. For this analysis we used a weighted interval score (WIS)[16], aggregated over the forecast horizon:

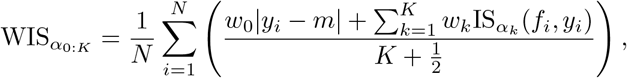

where *K* is the number of quantiles being evaluated, *w*_0_ is the weight given to the median and m is the predictive median. As above, *f*_*i*_ and *y*_*i*_ are the projected and observed data respectively on day *i*. IS_α_(*f*_*i*_, *y*_*i*_) is the interval score on day *i*, given by:

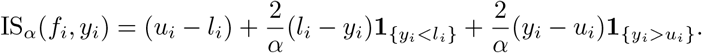

Here, 1 is the indicator function, meaning that 1_{*yi<li*}_ = 1 if *y*_*i*_ < *l*_*i*_ and 0 otherwise. The terms *l*_*i*_ and *u*_*i*_ denote the *α*/2 and 1 − *α*/2 quantiles of *f*_*i*_. The first term in the interval score gives the sharpness of the forecast, and the latter two are penalty terms for observations falling below *l*_*i*_ and above *u*_*i*_ respectively. For this analysis, we set *w*_0_ = 1/2 and *w*_k_ = *α*_*k*_ /2 as suggested in the literature[16]. The use of the WIS was chosen as, unlike the MAE, it takes into account the confidence intervals as well as the central estimates of the projections, and can give us values for the score at each time step. A projection will receive a good (small) WIS score if central estimates sit close to the observed data and confidence intervals surrounding the central estimate are both narrow and cover all the true data. We calculated the WIS for each time step of the MTPs, and took the mean to get an overall score of the projection similarly to the MAE.

#### 2.4.3 WIS on the log scale

We also calculated the WIS of the log transformed data, by taking the natural logarithm of the both the recorded data and forecasts before scoring. This can be seen as an approximation of a probabilistic counterpart to the symmetric absolute percentage error (SAPE), by considering the absolute error of the log transformed data, ϵ^∗^:

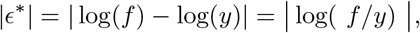

using the Taylor expansion:

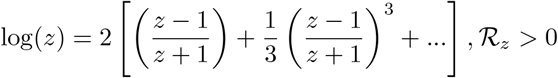

and, assuming that *f* ≈ *y*, we can approximate the absolute error, |ϵ^∗^|:

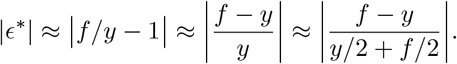

The alignment with SAPE has been shown to hold reasonably well even if the predicted and observed value differ by a factor of 2 or 3 [27]. In the rest of this paper we will refer to the WIS of the log transformed data as log WIS, for brevity.

#### 2.4.4 Empirical Coverage

For a forecast horizon, *h*, and projection interval width, 1 − *α*, the empirical coverage of a model (often referred to also as calibration[28]) is calculated as the proportion of forecast targets (across all forecast dates) for which the projection interval contained the true value; a well calibrated model has empirical coverage equal to the width of the nominal projection interval (i.e. the 50% projection interval should contain the true value 50% of the time). We calculated the empirical coverage for the 50% and 90% projection intervals over a range of forecast horizons from 1 to 21 days.

#### 2.4.5 Sharpness

Sharpness measures how good a model is at producing narrow (sharp) projection intervals. We measured sharpness as the weighted sum of the width of the 50% and 90% projection intervals, choosing weights of *w*_*k*_ = α_*k*_ /2 as we did for the WIS calculation, and again aggregating over the forecast horizon:

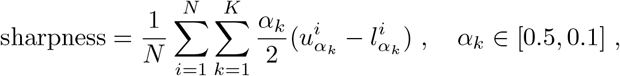

where *K* is the number of quantiles being evaluated,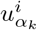and 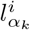 are the upper and lower bounds respectively for a given quantile *α*_*k*_ on day i. We calculated the sharpness for all the models, projected out to a 28 day forecast horizon, against the publishing date. It is worth noting that sharpness is a property of the forecast only. Therefore, we evaluated the predictive performance based on “the paradigm of maximizing the sharpness of the predictive distributions *subject to calibration*“[29], and took the calibration to be directly evaluated by empirical coverage.

#### 2.4.6 Bias

Bias measures a model’s tendency to over or under predict. To calculate this we simply subtracted the sum of recorded observations from the sum of the central model estimate for a given metric, and did this for each publishing date.

### 2.5 Forecast comparison

In order to evaluate the utility of the ensemble approach, we compared the predictive performance of the combined MTPs with the performance of the constituent models. We measured the performance by calculating the MAE, WIS, sharpness and empirical coverage of the combined model and its constituents over a range of publication dates and forecast horizons for the three metrics. We did this for MTPs published between November 2021 and December 2022. For the empirical coverage we aggregated the models over the entire time period, and evaluated over a range of forecast horizons. For WIS, sharpness and MAE we evaluated and compared all of the models at 7, 14, and where applicable 21 and 28 day forecast horizons for each published MTP over the range of publishing dates.

## 3 Results

### 3.1 COVID-19 hospital admissions, bed occupancy and deaths data between November 2021 - December 2022

Each of the models were able to produce time series for at least one of hospital admissions, bed occupancy and deaths related to COVID-19. These time series are shown in fig. 1 A, C and E. Hospital admissions, bed occupancy and deaths were all relatively low at the tail end of 2021, reaching a 7 day average trough of 643 new hospital admissions, 5,900 beds occupied and 95 deaths on 25 November, 4 and 10 December respectively. At this time, the Delta (B.1.617.2) variant was still responsible for the vast majority of cases [30]. In late November 2021, the first Omicron cases were detected in the UK, onsetting the first Omicron “wave” which saw infections increase before peaking with a 7 day average of 2,040 new hospital admissions, 16,696 beds occupied and 253 deaths on 1, 12 and 19 January 2022 respectively. This wave consisted of a mix of BA1.1 and B1.1.529, visible in fig. 1G, which shows the relative proportions of the variants over time. All legal COVID restrictions were officially lifted in England on 24 February 2022. In late March, booster vaccinations were offered by NHS England to people aged over 75 and anyone over the age of 12 who was considered medically vulnerable. Around the same time the BA.2 Omicron sub-lineage led to the second Omicron wave in spring and early summer, which peaked with 2,116 new hospital admissions, 16,600 beds occupied and 250 deaths on 28 March, 7 and 10 April respectively . On 1 July, the UK government dashboard moved from daily to weekly reporting [31]. The BA.4 and BA.5 sub-lineages co-existed throughout autumn 2022, as shown in fig. 1, which helped drive the third Omicron wave that caused 7 day averages of 1,864 new hospital admissions, 13,849 beds occupied and 189 deaths on 10, 16 and 18 July respectively. The wave at the end of the year consisted of a mixture of previously established Omicron sublineages, namely BA.2, BA.4 and BA.5. This wave peaked with 7 day average counts of 1,212 new hospital admissions, 10,560 hospital beds occupied and 152 deaths on 4, 15 and 19 October respectively.

**Figure 1:**
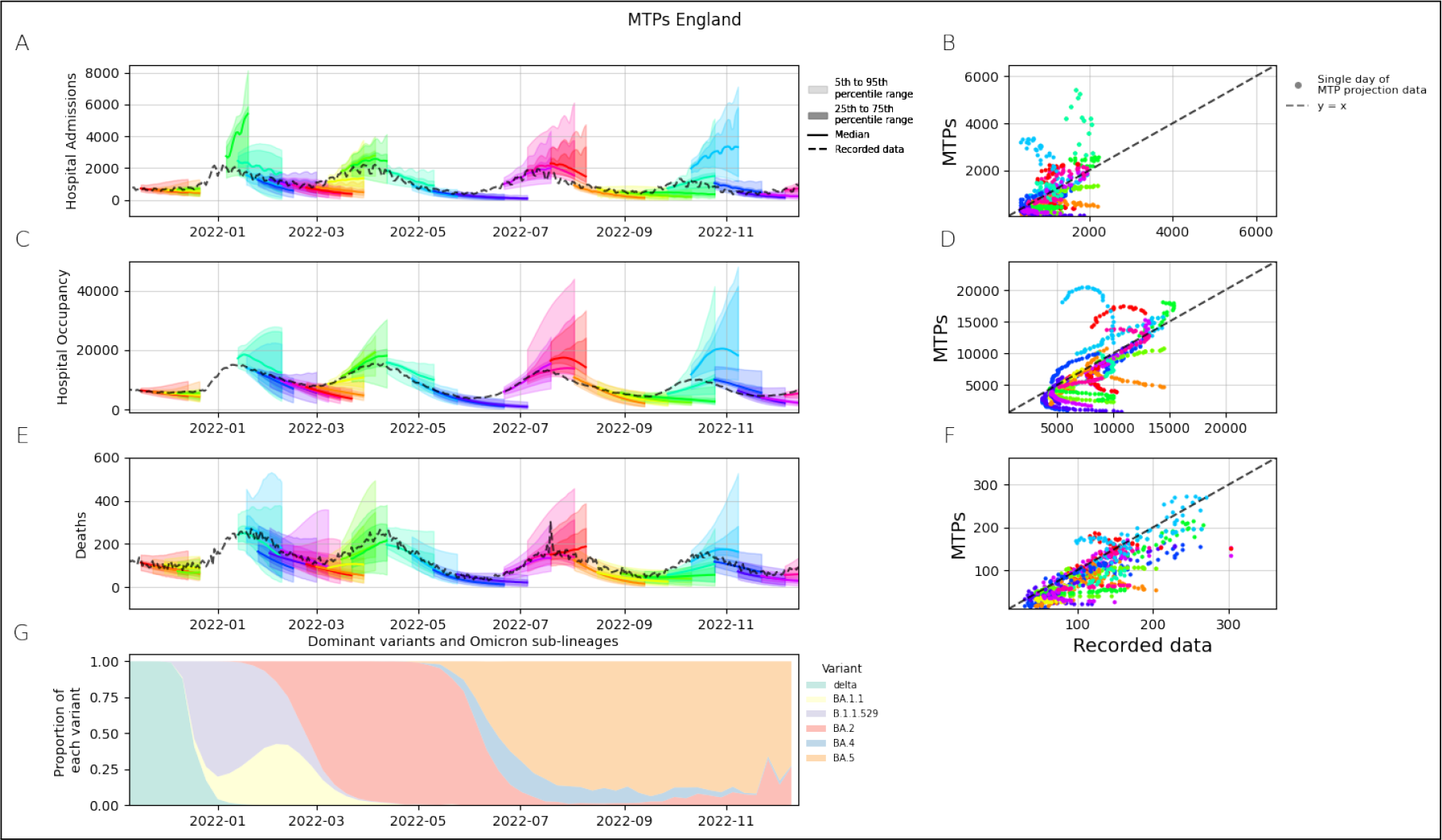
Combined MTPs for hospital admissions, hospital bed occupancy and deaths plotted against the recorded data in plots A, C and E respectively, for the period between November 2021 to December 2022. Hospital admissions was the only metric for which a combination was published on 06-01-2022, meaning there is one extra combined MTP shown in plot A. Plots B, D and F show scatter plots of the forecast hospital admissions, bed occupancy and deaths plotted against the recorded data, where each point represents a single day of data. Each colour in plots A - F represents a single published combined forecast, each with a forecast horizon of 28 days. Plot G uses data from [30] and shows the relative proportions of cases belonging to each variant and subvariant from the genomes that were sequenced.

### 3.2 Evolution of Medium-Term Projections

We gain a qualitative understanding of the overall model performance by observing the plots of the model’s projections against the recorded data. The combined model plots are given in fig. 1. The left hand column shows how the MTPs evolve over time, with each colour in the plot representing a specific publishing date. The right hand column shows the central estimate of the combined model plotted against the observed data, with a *y* = *x* line for comparison. Each point in the plot represents the projected value for each time step of the projected data. A perfect fit to the data would be along the *y* = *x* line in this plot. Therefore, the perpendicular distance of points from this line show the discrepancy between the published MTPs’ central estimate and the recorded data.

From fig. 1, we can see that overall the combined MTPs were in good agreement with the data for most of the Omicron epidemic waves. The MTPs were able to predict the trends of increasing and declining epidemic curves, although the visual inspection suggests that the combined model generally performed worse when forecasting hospital bed occupancy during the Omicron sub-lineage waves in the summer and autumn of 2022. The trend is less clear for hospital admissions and deaths. The 90% confidence intervals are widest when projecting over the epidemic peaks. Figure 1G shows a stacked area plot of the proportions of each variant over time from genomic sequencing data[30] ^2^.

### 3.3 Forecast Evaluation

#### 3.3.1 Combination

The combined model performed worst during the first Omicron (B.A.1) wave in January 2022. Both the mean WIS and the MAE reached their maximum on 5 January for hospital admissions, and on 12 January for hospital occupancy and deaths^3^. Furthermore, the models were only projected out to 2 weeks in this instance^4^, so we do not have results for forecast horizons beyond 14 days. In general, the model ensemble performs better when the number of hospital admissions is either nearing or just past an epidemic peak, and has the worst MAE and WIS around the time of the epidemic wave peak. Unsurprisingly the combined model generally has a much lower MAE for the 0 - 7 day forecast window compared with the 7 + days forecast windows, with the notable exceptions occurring when hospital admissions had just passed the epidemic wave peak, in early January 2022 and mid July 2022. The log WIS is generally higher for both peaks and troughs of the epidemic, suggesting a worse performance around turning points in general, a result which is “hidden” in the naturaly scaled scoring method. The log WIS from May 2022 to July 2022 also has a greater dependence on the forecast window than the natural scale score, with the performance being much better for the forecast windows closer to the start date. An evaluation of the model performance for hospital admissions is plotted in fig. 2, and similar plots are given for hospital bed occupancy and deaths in the supplementary material.

**Figure 2:**
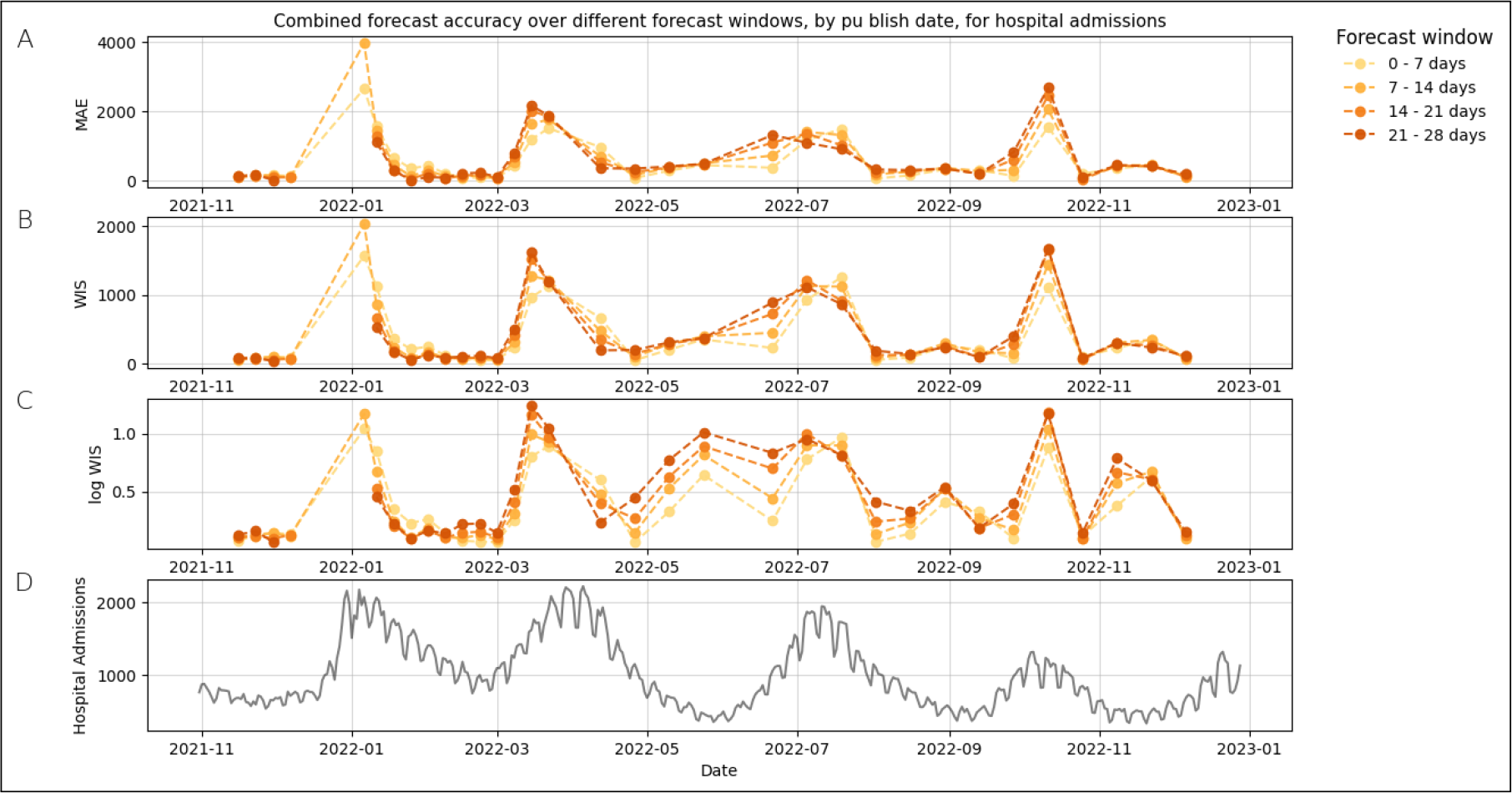
Mean absolute error (MAE) and Weighted interval score (WIS) of the Combined MTPs, for hospital admissions, during the period between November 2021 to December 2022. The different colours represent different parts of the forecasting window up to 4 weeks. In some instances a combined projection wasn’t published beyond 2 weeks (i.e. in January 2022), hence there are some publishing dates which do not have plotted values for longer forecast horizon windows. Plot A shows the MAE for the naturally scaled data, and plots B and C show the WIS for the natural and logarithmic scale respectively. The observed data is shown in plot D for reference.

#### 3.3.2 Individual Models

We found that the predictive ability of each individual model changed over time, and whilst the WIS of the combination was often not the lowest, no single model consistently outperformed the combination. The DDMs performed better for shorter forecast horizons than longer ones, and generally performed worse than the mechanistic models (PBMs and ABMs) during epidemic peaks (fig. 3). This is most notable in July 2022 during the Omicron B.A.5 peak, for which the two worst performing models were DDMs. The mechanistic models were better than the DDMs at predicting epidemic peaks, but performed worse when approaching a trough. The individual models are heterogeneous in their predictive ability (fig. 3), however, the combination smooths out many of these heterogeneities, and is therefore more robust to changes in the status of the epidemic than any of the individual models. The log WIS shows the benefit of a model combination more clearly around the epidemic troughs when compared with the natural scale. In the periods from February 2022 to March 2022 and from May 2022 to July 2022 the combined model has a log WIS of nearly half that of the worst performing models (fig. 3).

**Figure 3:**
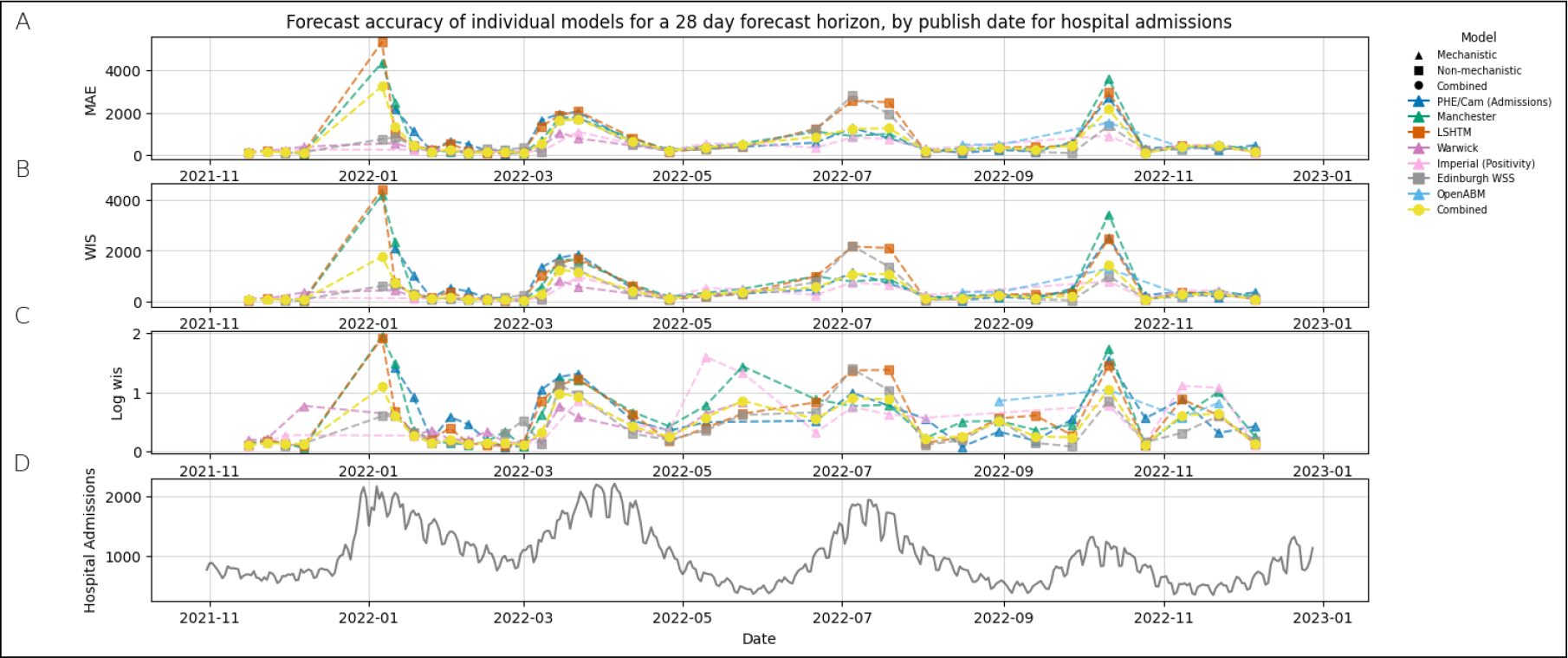
Mean absolute error (MAE) and Weighted interval score (WIS) of the individual models’ submitted MTPs for hospital admissions for the period between November 2021 to December 2022. The MAE and WIS were calculated with a forecast horizon of 28 days. Plot A shows the MAE for the naturally scaled data, and plots B and C show the WIS for the natural and logarithmic scale respectively.

#### 3.3.3 Empirical coverage, sharpness and bias

The combined model performs the best overall at estimating both the 90% and 50% confidence intervals compared to any single model, albeit with the EpiNow2 model being marginally superior for the 90% confidence interval over very short forecast horizons (< 4 days) (fig. 4A and B). For hospital admissions, in both cases the empirical coverage is closest to the target around the 7 day forecast horizon. The combined model is one of the least sharp compared with the individual models across the majority of publishing dates (fig. 4C). However, the models which are consistently the “sharpest” for hospital admissions, (i.e. Manchester, Imperial and PHE/Cam (admissions)) also have the lowest empirical coverage.

**Figure 4:**
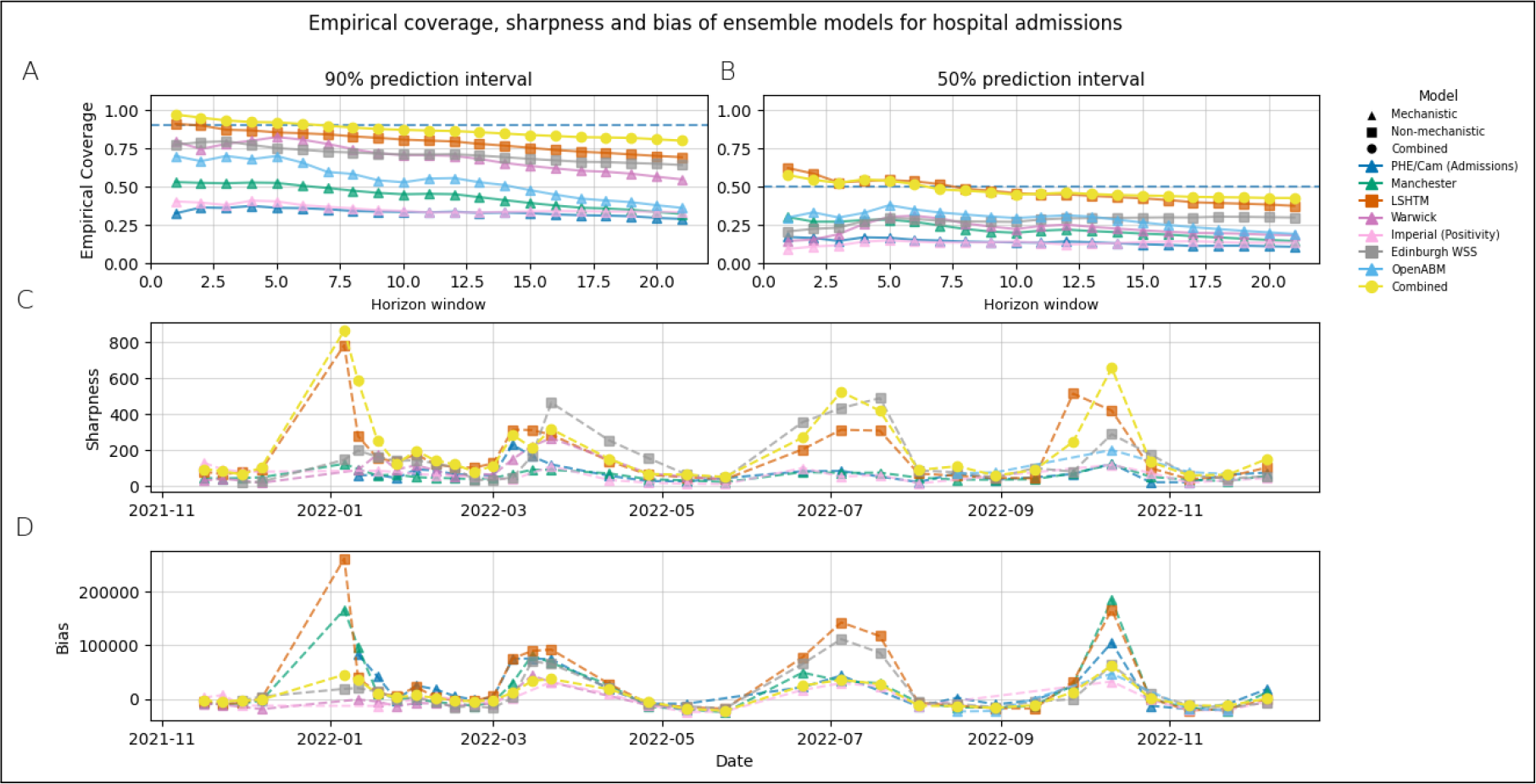
Plots A and B show the empirical coverage of the individual and combined models for forecasting windows of 1 - 21 days for the 50% and 90% confidence intervals. Sharpness and bias are shown in plots C and D, averaged over a forecast horizon of 28 days. The plots shown are for hospital admissions only.

All of the models in the ensemble have a tendency to over predict, demonstrated by the bias being positive far more often than negative (fig. 4D). This effect is greatest around the epidemic peaks. Some underprojection does occur at the troughs, but the effect is much smaller.

Equivalent plots for hospital bed occupancy and deaths are shown in the supplementary material. For occupancy, the empirical coverage of the combined model is over-estimated for the 90% confidence interval at forecast horizons lower than 12 days, but the combination has the best empirical coverage for forecast horizons beyond this. For the 50% confidence interval the combined model has the best empirical coverage beyond 7 day forecast horizons. For deaths the empirical coverage is overestimated by the combined model as, for both 50% and 90% confidence intervals, more than 50% and 90% of the recorded data points are covered respectively. Despite this, the combination still has a better empirical coverage than any single model in the ensemble.

## 4 Discussion

This paper showcases the process of ensemble modelling of the Omicron epidemic waves over the period November 2021-December 2022. We illustrated how medium term projections (MTPs) can be derived from a model ensemble with three groups of models: PBMs, ABMs and DDMs. Additionally, we evaluated the probabilistic accuracy of combined and individual medium-term projections (MTPs) over this period.

Our results suggest that the ensemble model generated MTPs were in general the most accurate future projections (as measured by the mean MAE and WIS metrics) across different forecasts windows and time points, as they were able to buffer and overcome the variable accuracy of individual models.

The ensemble MTPs were aligned with the data for most of the study period, with this alignment better (as measured by the MAE and WIS scores) over periods of exponential increase or decline in the epidemic than around the epidemic peaks or troughs. The individual mechanistic models were better in projecting forward the timing and size of the epidemic peak, compared to both DDMs or the model ensemble. This is not a surprise; mechanistic models include specific mechanisms, such as the depletion of the susceptible pool, that are an important aspect of correctly capturing the pandemic peak characteristics.

Ensemble modelling, although common practise in climate science [3, 4, 5], is a developing area in epidemiological modelling. The approach in epidemiological modelling in the pre-COVID-19 era was often focused on developing a specific disease or intervention question, albeit with notable modelling efforts present in HIV epidemiology and in the response to the Ebola outbreak [13, 14]. During the COVID-19 epidemic, with the vast popularisation of modelling, a large number of models were being developed using similar assumptions and were parameterised and/or calibrated to the same or similar data sources. This therefore lent itself to ensemble modelling. Our findings support the notion that variability emerging from different model parameters, assumptions and structures can be reduced by using a combined estimate, which characterises the overall uncertainty in a system better than any single model, and that a model ensemble produces more accurate forward projections.

### 4.1 Strengths of our work

One of the strengths of our approach is that we use a variety of models: those that use one type of data or a number of data streams or those that include mechanisms or not. While the ensemble models can be broadly stratified into the three structure-based groups of PBMs, ABMs and DDMs, they are all technically autonomous and have been continuously developed over the pandemic period. Furthermore, we note that although models have aimed to include similar assumptions, there are subtle differences between them. For example, models such as EpiNow2 are data driven and do not explicitly model mechanisms such as waning immunity or the depletion of the susceptible pool, but incorporate their effect when fitting to data. Mechanistic models such as Imperial’s sir-covid or PHE/Cambridge, explicitly model the mechanisms of disease spread, and included certain interventions such as vaccination rollouts and school term times. Having these different models in the ensemble not only enriches the variety of outcomes, but also adds wider uncertainty to the combined model projections.

Our results for model sharpness and empirical coverage show that the use of a model ensemble adds uncertainty to the forward looking projections, whilst generally containing more data points within the 50% and 90% confidence intervals than any single model. In epidemiological models, the structure of the model and associated assumptions alongside the model parameters determine the characteristics of the resulting projections. For example, the Manchester model requires the modeller to decide on “change points” where the behaviour of viral transmission is changing, achieved by adding a new value for the *β* parameter which controls the transmission rate. Therefore if an epidemic trough is approaching in the near future due in part to a waning of immunity in the population, this model is less likely to capture the turning point as it will continue to project the current trend downwards more than another model which explicitly includes waning immunity as a parameter. Equally, the inherent uncertainty in parameters like waning immunity means that two models are highly unlikely to display the same behaviour even if they both model the mechanism directly. This is what adds uncertainty to the model ensemble and allows a wider window of possible outcomes to be generated than if a single model was used. Exemplifying this is the fact that, as mentioned in the previous section, the sharpest models in the ensemble have the lowest empirical coverage. So despite narrow projections, the confidence intervals of these models do not cover the data points as well as the combination. When aiding policy decisions, it is better to have an understanding of the possible futures, and the likelihood associated with each one, rather than have a very confident projection which does not contain the observed data appropriately within its confidence intervals[32].

Another strength of our model ensemble is that we use models that are calibrated to a variety of data sources, detailed in table A1. Having models that fit to a variety of data again widens uncertainties in the combined model projections. Furthermore, no data is free of bias so using multiple data sources reduces the bias associated with any single data source. For example, case data is highly sensitive to ascertainment biases, the scale of which can vary over time. Therefore, models that fit to case counts or positivity must be interpreted in the context of testing behaviours and policies at the time. However, admissions data is not free from bias either. The likelihood of being admitted to hospital varies greatly by age. Hence, without age-stratification in the model, it is likely that community transmission is under-estimated among younger age groups. Furthermore, the delay between being infected with COVID-19 and being admitted to hospital was on average far greater than that between infection and receiving a positive test, particularly at the time when free tests were readily available. This presents difficulties when trying to produce timely estimates of community transmission.

### 4.2 Limitations of this work

Our work has some limitations. Firstly, the model ensemble, which was used operationally, was not consistent over the full time period. Models in the ensemble were being developed continuously, and therefore were subject to ongoing changes. For those models built and maintained by members of SPI-M-O, it was not possible to track all of these changes. Furthermore, the number of models in the ensemble would change for various reasons. This is shown clearly in fig. 3, where the OpenABM model is only included in the projections between September 2022 - November 2022. The constantly evolving ensemble makes it more difficult to assess the performance over the time period.

Secondly, the models in the ensemble were combined using equal weights stacking. It was beyond the scope of this work to explore alternatively weighted combination methods, but this is something we are planning on undertaking in the future. Specifically, future work would focus on a subset of the models over a shorter period to enable a more even comparison, as well as using an alternative combination and weighting strategies. In order to be operationally viable an alternative weighting would, however, need to be adaptive, in order to incorporate different models entering or leaving the ensemble.

Finally, none of the models in the ensemble picked up on the oscillatory nature of the epidemic (see fig. 1). This is to be expected as the models were designed for the medium term of 4 - 6 weeks, and the oscillation is a longer term trend with a period of roughly 10 weeks. Future work could therefore look to combine medium term forecasts similar to those discussed in this paper, with longer term pattern matching or ARIMA type models, in order to try and more accurately capture the oscillations of the epidemic in its later stages.

### 4.3 Conclusions

In summary, our results illustrate that the combined MTPs, produced from an ensemble of heterogeneous epidemiological models across different Omicron epidemic waves, were a closer fit to the data than the individual models throughout late 2021 and during 2022. The alignment with the data was best during the periods of epidemic growth or decline, with the 90% confidence intervals widest around the epidemic peaks. Combined MTPs also improve the robustness and reduce the bias associated with an individual model projection. Hence, we advocate development of formal national and international ensemble modelling hubs for infectious disease modelling as a key step in preparing for the next outbreak or pandemic.

## 5 Data and code availability

The data and code for the analyses undertaken will be available from the corresponding author upon a reasonable request.

Each model output included in this paper was a joint effort of many people in the various modelling teams. In order to stick to the maximum number of co-authors allowed by the journal, a decision was made to only include one representative from each modelling team as co-author on this paper. This representative submitted the model outputs, attended and discussed these at the weekly SPI-M-O/ UKHSA meeting, representing the larger efforts by their team. We would therefore like to acknowledge the huge amount of work done by all contributors within various modelling teams, as this work would not have been possible without them. These contributors include, but are not limited to: Veronica Bowman, Thomas Maishman, Thomas House, Katrina Lythgoe, Neil Ferguson, Robert Hinch, Christophe Fraser, John Edmunds, David J Wallace and James A Ackland.

## Data Availability

The data and code used in this analysis can be available from the corresponding author upon publication and upon a reasonable request.

## A Model and data descriptions

**Table A1.**
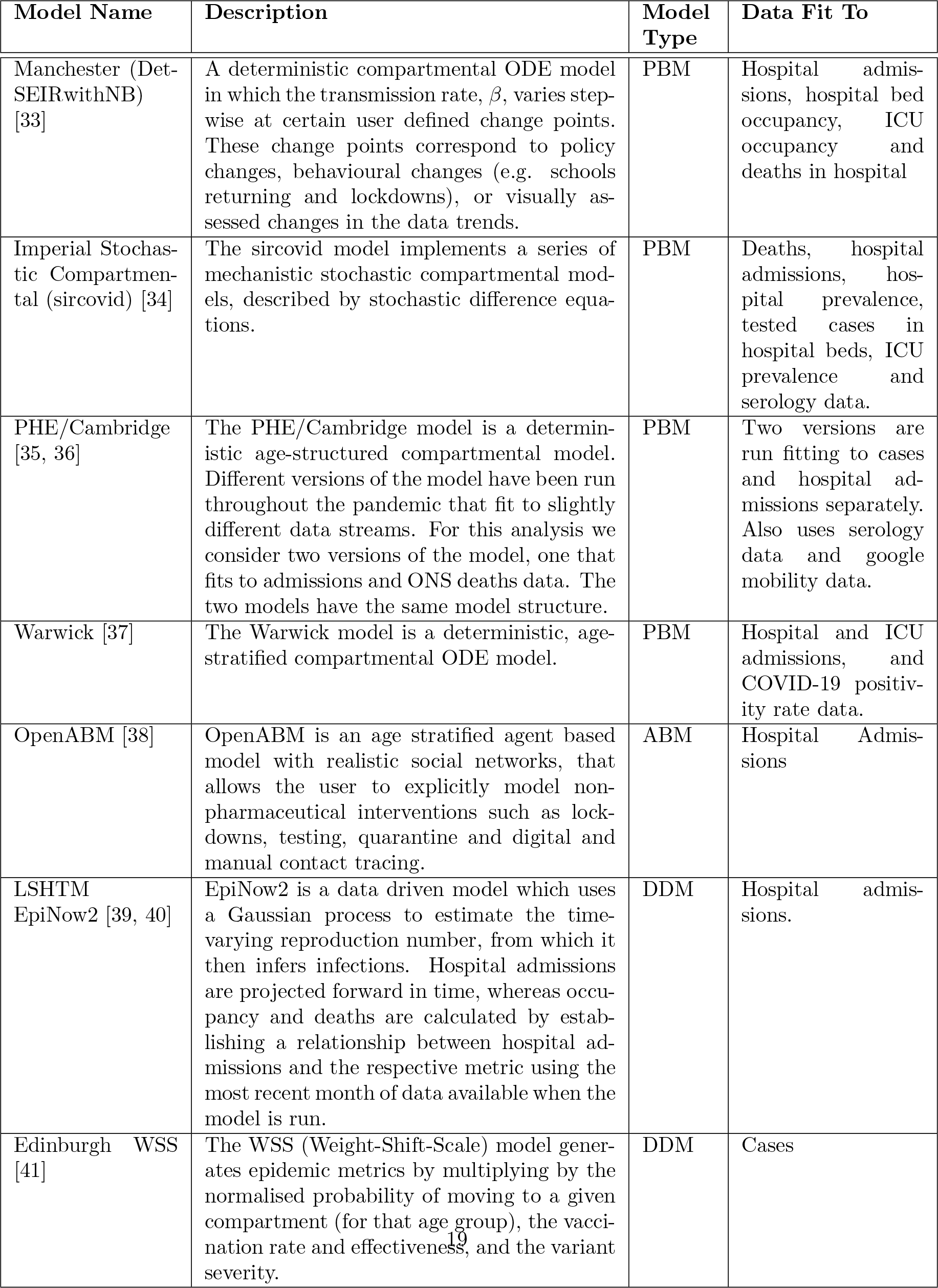
: Outline of the epidemiological models used to generate MTPs for the English COVID-19 epidemic. We list the names of the models, as well as their main modelling characteristics and the data to which they are calibrated against.

## Footnotes

1 The standard consensus projection length was either 4 or 6 weeks. However, at times of significant uncertainty, for example in the early stages of the Omicron variant emerging, projections were published with shorter forecast horizons.

2 The genomics data “only includes a subset of UK SARS-CoV-2 sequencing surveillance data and should not be used to estimate frequency of SARS-CoV-2 variants circulating” [30], however it still provides a good estimate of the relative proportion of different variants, and the rough time that each becomes dominant.

3 There was no published combination of projections on 5 January for these metrics.

4 see footnote on page 3

