## Supplemental material for "Combining models to generate consensus medium-term projections of hospital admissions, occupancy and deaths relating to COVID-19 in England"

### S1 Additional time series plots for hospital bed occupancy and deaths

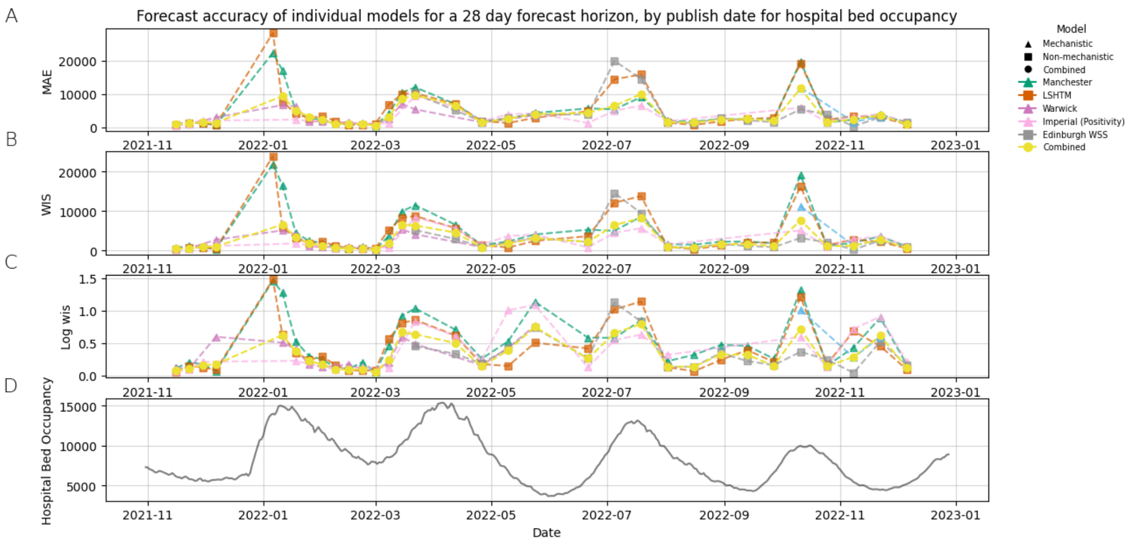


**Figure S1:** Mean absolute error (MAE) and Weighted interval score (WIS) of the individual models' submitted MTPs for hospital bed occupancy for the period between November 2021 to December 2022. The MAE and WIS were calculated with a forecast horizon of 28 days. Plot A shows the MAE for the naturally scaled data, and plots B and C show the WIS for the natural and logarithmic scale respectively. The observed data is shown in plot D for reference.

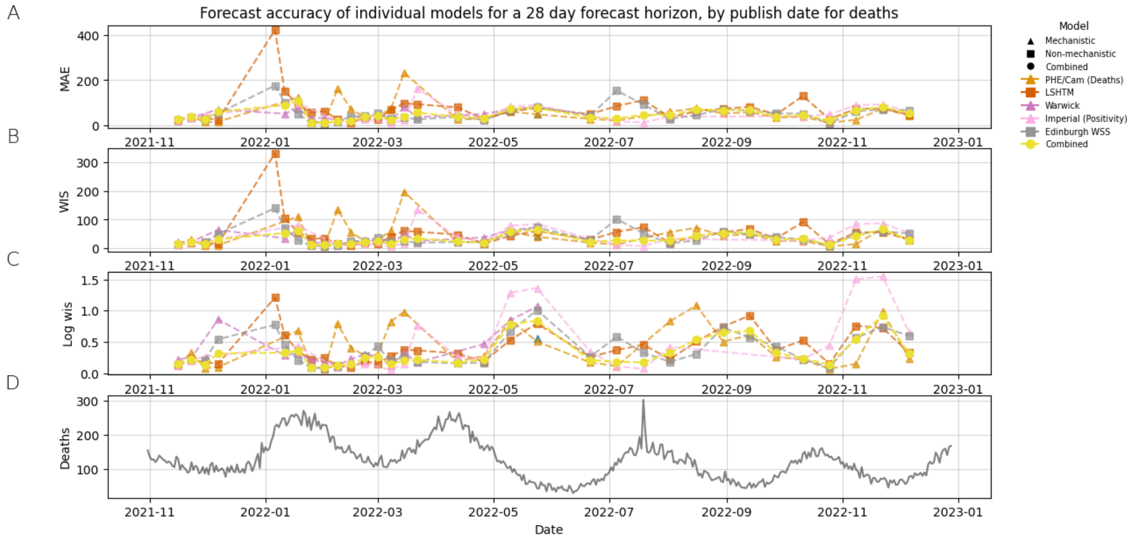


**Figure S2:** Mean absolute error (MAE) and Weighted interval score (WIS) of the individual models' submitted MTPs for deaths within 28 days of a positive test, for the period between November 2021 to December 2022. The MAE and WIS were calculated with a forecast horizon of 28 days. Plot A shows the MAE for the naturally scaled data, and plots B and C show the WIS for the natural and logarithmic scale respectively. The observed data is shown in plot D for reference.

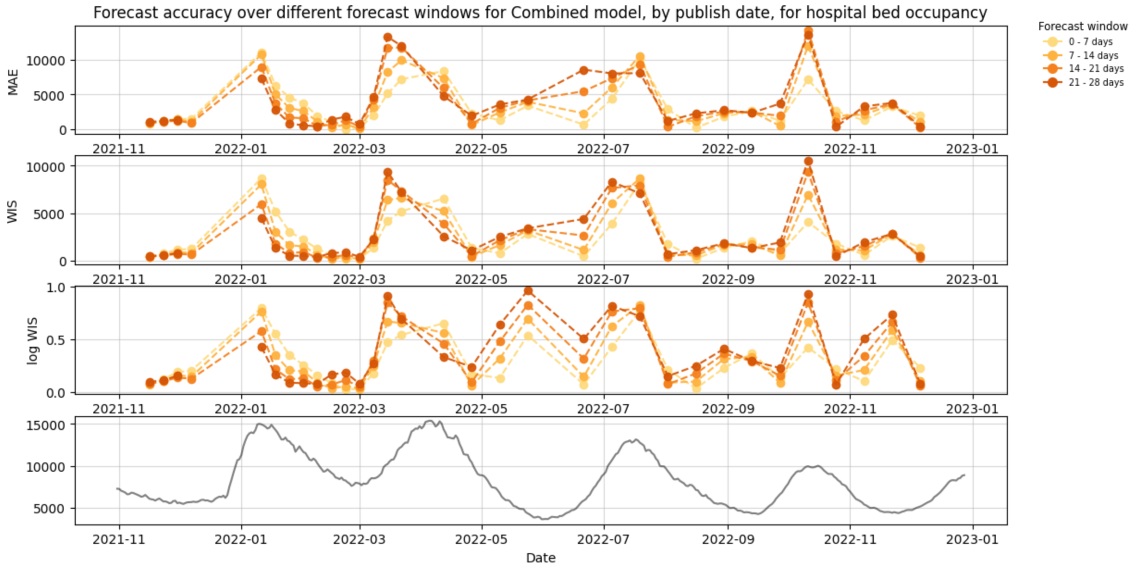


**Figure S3:** Mean absolute error (MAE) and Weighted interval score (WIS) of the combined MTPs, for hospital bed occupancy, during the period between November 2021 to December 2022. The different colours represent different parts of the forecasting window up to 4 weeks. Plot A shows the MAE for the naturally scaled data, and plots B and C show the WIS for the natural and logarithmic scale respectively. The observed data is shown in plot D for reference.

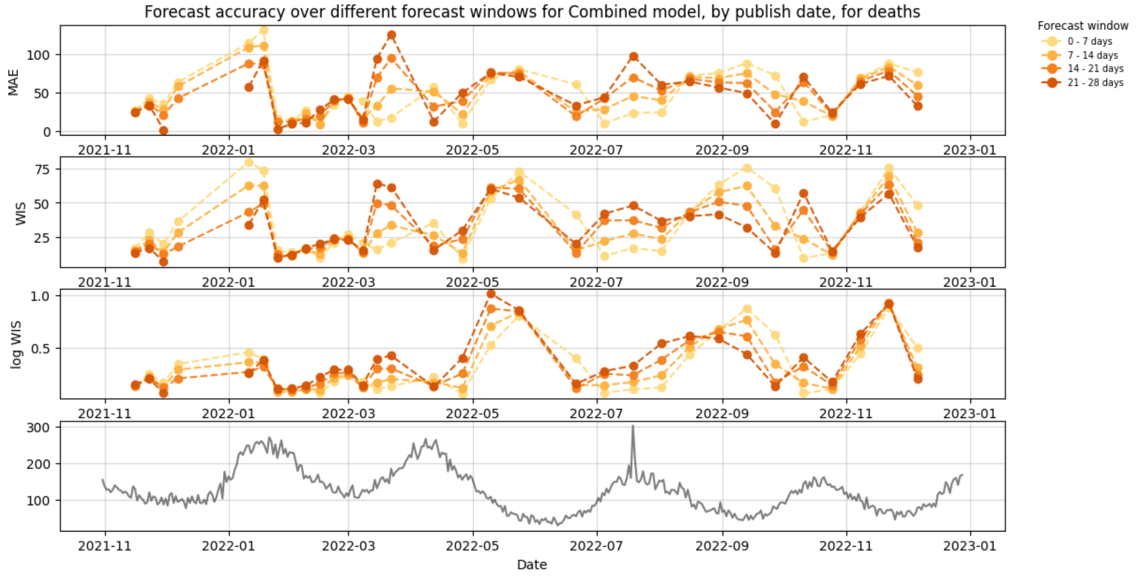


**Figure S4:** Mean absolute error (MAE) and Weighted interval score (WIS) of the combined MTPs, for deaths within 28 days of a positive test, during the period between November 2021 to December 2022. The different colours represent different parts of the forecasting window up to 4 weeks. Plot A shows the MAE for the naturally scaled data, and plots B and C show the WIS for the natural and logarithmic scale respectively. The observed data is shown in plot D for reference.

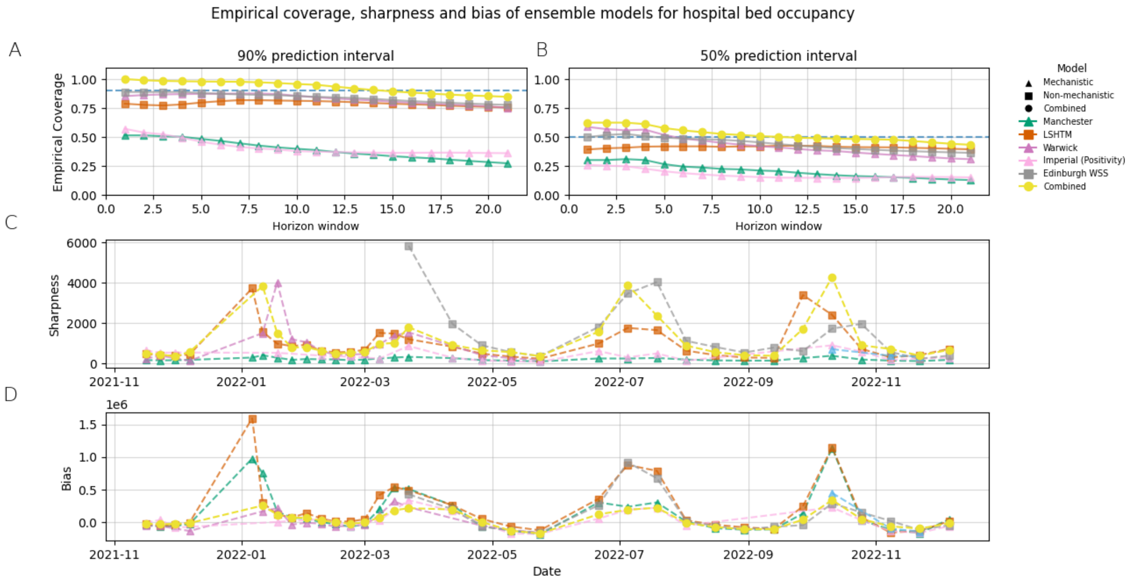


**Figure S5:** Plots A and B show the empirical coverage of the individual and combined models for forecasting windows of 1 - 21 days for the 50% and 90% confidence intervals. Sharpness and bias are shown in plots C and D, averaged over a forecast horizon of 28 days. The plots shown are for hospital bed occupancy.

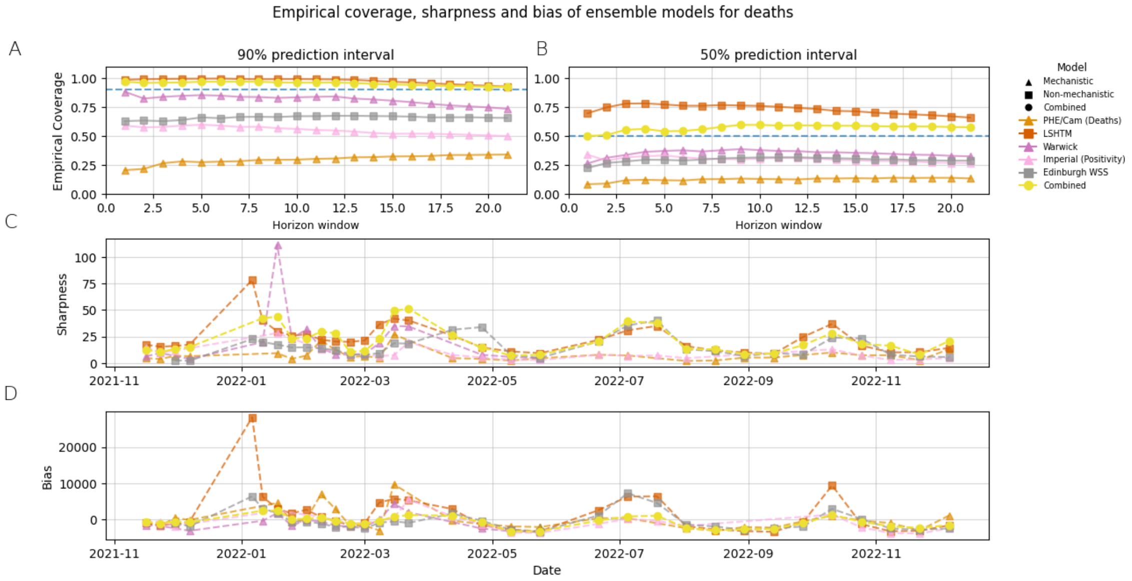


**Figure S6:** Plots A and B show the empirical coverage of the individual and combined models for forecasting windows of 1 - 21 days for the 50% and 90% confidence intervals. Sharpness and bias are shown in plots C and D, averaged over a forecast horizon of 28 days. The plots shown are for deaths within 28 days of a positive test.
